# BioDecoder: A miRNA Bio-interpretable Neural Network Model for Noninvasive Diagnosis of Breast Cancer

**DOI:** 10.1101/2023.01.31.23285308

**Authors:** Lei Liu, Weili Lin, Suqi Cao, Liu Yang, Sheng Gao, Na Jiao, Lixin Zhu, Ruixin Zhu, Dingfeng Wu

## Abstract

Early diagnosis of breast cancer remains a major clinical challenge. Liquid biopsy has become a powerful tool for cancer diagnosis by the aid of various the state-of-the-art detection technologies and artificial intelligence (AI) methods. Although the prediction performance is superior, the clinical application of existing AI models is greatly limited due to their poor interpretability. Here, we designed a miRNA-Gene-Module-Pathway-Disease biological decoding path, and developed BioDecoder thereof, a miRNA bio-interpretable neural network model for breast cancer early diagnosis. We demonstrated that BioDecoder could achieve early non-invasive diagnosis of breast cancer with a remarkable performance (AUC = 0.989) and showed strong generalizability in an external cohort (AUC = 0.890). Meanwhile, the biologically interpretable results of BioDecoder revealed that significant changes in metabolic pathway and oxidative phosphorylation were the main action pathways of circulating miRNA in breast cancer. Our study indicates that BioDecoder offers the promise of non-invasive early diagnosis of breast cancer and can be generalized to other cancers and corresponding biomarkers.

## Introduction

Breast cancer is one of the most common malignancies worldwide. According to the data released by GLOBOCAN 2020, female breast cancer has surpassed lung cancer as the most commonly diagnosed cancer with about 2.3 million (11.7%) new cases in 2020, being the leading cause of cancer mortality among women ^1^. Published research has revealed that the 5-year average survival rate of in situ female breast cancer reaches 99.0%, while those of regional- and distant-stage breast cancer are only 86.0% and 29.0%, respectively ^2^. Lokong et al. ^3^ also reported that delayed diagnosis was an important reason for the higher breast cancer mortality in low-income countries. Therefore, early screening and diagnosis are essential to improve the overall survival rate of breast cancer. Tissue biopsy is the gold standard for clinical diagnosis of breast cancer; however, as an invasive test it is not suitable for early detection ^4^. Currently, mammogram screening has been commonly used for early diagnosis of breast cancer but with risks of overdiagnosis and radiation exposure ^5-7^. Hence, it is imperative to develop an accurate and non-invasive alternative tool for the early detection of breast cancer.

Liquid biopsy has become an important means of clinical early screening of cancer^8,9^. It can detect and analyze circulating tumor DNA (ctDNA), RNA (i.e., mRNA, miRNA), circulating tumor cells (CTC), and exosomes in plasma, urine, and other body fluids, providing information that is difficult to capture in medical imaging^10, 11^. Compared with tissue biopsy, liquid biopsy is non-invasive and easier to monitor tumor oncogenesis, metastasis and treatment response in real time ^12, 13^. Although the diagnosis of breast cancer is challenging due to heterogeneity ^14^, circulating carcinoma proteins, circulating tumor cells, ctDNA, circulating miRNA, and other biomarkers have been applied in liquid biopsy research of breast cancer and achieved good predictive performance ^15, 16^. Among them, the circulating miRNA plays an important role in tumor pathogenesis as oncogenes or tumor suppressors ^12, 17^, making it a promising biomarker for breast cancer diagnosis. In previous research, using machine learning algorithms, a panel of five miRNAs (miR-1246, miR-1307-3p, miR-4634, miR-6861-5p and miR-6875-5p) was demonstrated to detect breast cancer with 89.7% accuracy ^18^, and another set of seven miRNAs including has-miR-126-5p and has-miR-144-3p showed predictive power for triple-negative breast cancer with an area under the receiver operating characteristic curve (AUC) of 0.814 ^19^.

Artificial intelligence (AI), including traditional machine learning algorithms and deep learning architectures, has greatly altered the research paradigm in medical science, and has brought new breakthroughs in precise diagnosis, treatment and prognosis of cancer ^20^. Relying on the development of AI, liquid biopsy has become a powerful tool for cancer diagnosis ^21^. The inherent black-box nature of most AI models, however, hinders their interpretability and widespread clinical application ^22^. To help alleviate this problem, eXplainable AI (XAI) ^23^ has been introduced. Research have revealed that feature importance, model perturbation, feature association, and prior knowledge, etc., can be utilized to improve the interpretability of AI models ^24, 25^. By integrating prior biological knowledge, bio-interpretable models (white-box solution) can be constructed to capture potential causality and uncover the underlying biological process of diseases with better model credibility and generalizability, thereby promoting the research of disease mechanisms and the identification of therapeutic targets. For example, a recent study by Elmarakeby et al. ^26^ have demonstrated the capacity of biological XAI model for revealing novel molecularly altered candidates and predicting the staging of prostate cancer patients. Consequently, development of a breast cancer early diagnostic biological XAI model promises great benefits for further popularizing the clinical application of breast cancer liquid biopsy.

This study was undertaken to design a miRNA biological decoding path (miBDP) and develop BioDecoder, a miRNA bio-interpretable neural network model, for breast cancer early screening and diagnosis. Integrating prior biological knowledge and AI technology, BioDecoder dramatically ameliorated its biological interpretation ability under the premise of ensuring prediction performance. The findings drawing from BioDecoder provide new insights into the pathogenesis and treatment of breast cancer.

## Results

A set of 4113 serum samples, including 2833 control samples (i.e., 2686 non-cancer samples, 93 prostate disease samples and 54 benign breast disease samples) and 1280 breast cancer samples, and corresponding profiles of 2540 circulating miRNAs were obtained as the discovery cohort (Table S1) ^18^. We developed a miRNA bio-interpretable neural network model (BioDecoder) to diagnose breast cancer, whose performance was compared with traditional black-box models (i.e., random forest [RF] and fully connected neural network [FCN]). The potential mechanism of miRNA in breast cancer was then explained through BioDecoder. Finally, the predictive performance of BioDecoder was validated on an external cohort (11 control samples and 122 breast cancer samples, Table S2) by transfer learning (Figure 1A).

**Figure 1.**
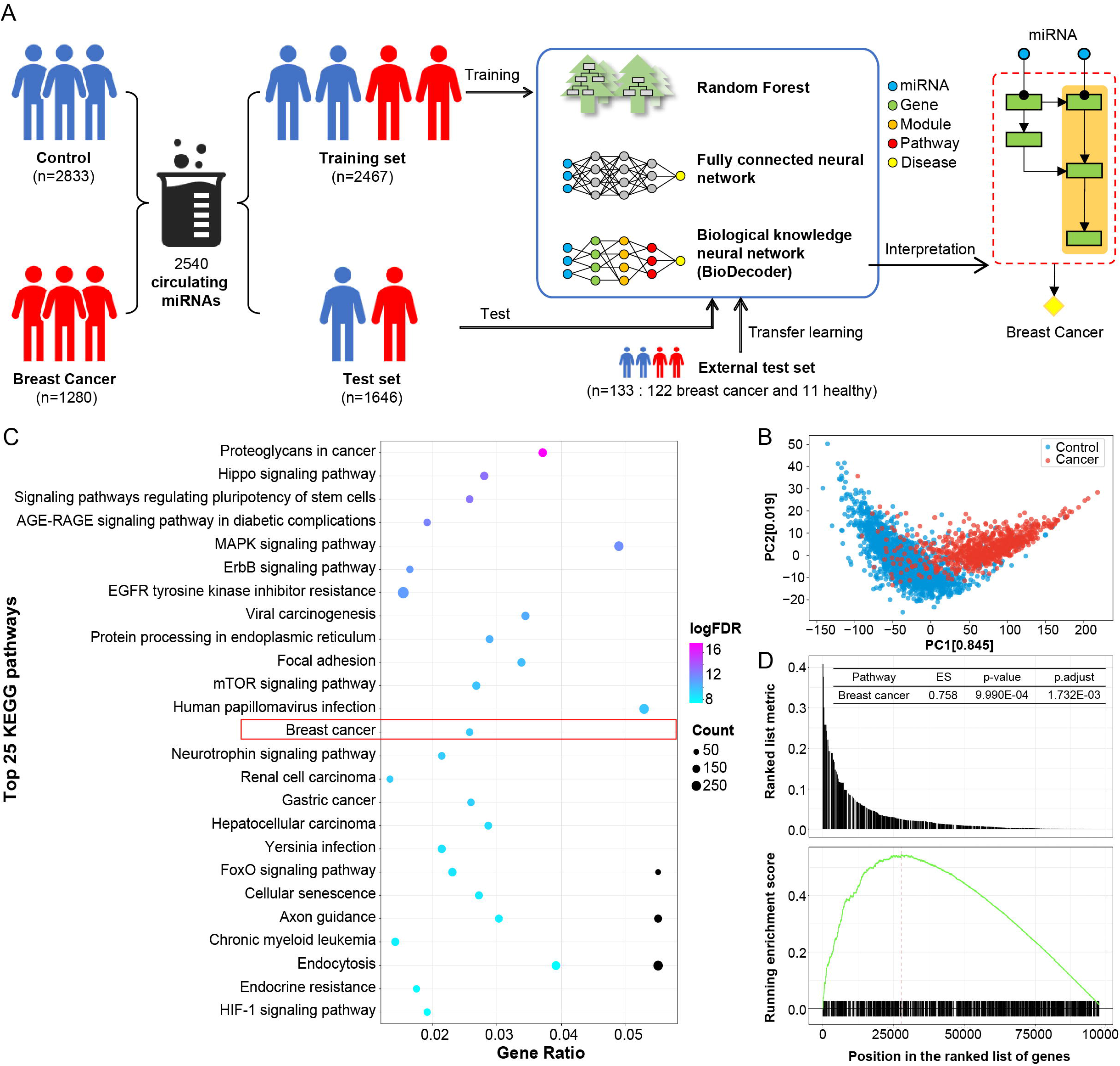
Differential changes of circulating miRNAs in breast cancer. (A) The workflow of this study for breast cancer diagnosis based on circulating miRNAs. (B) Principal component analysis (PCA) of miRNA profiles showed different distribution between breast cancer and control samples in the discovery cohort. (C) The top 25 KEGG pathways enriched by the target genes of differential miRNAs. (D) Gene set enrichment analysis results of differential miRNA target genes in breast cancer pathways.

### Differential circulating miRNAs as biomarkers for breast cancer diagnosis

Circulating miRNAs in serum have potential for breast cancer diagnosis ^27, 28^, which was confirmed in our discovery cohort (Figure 1B). Seven hundred and ten miRNAs with significant differences between breast cancer and control samples were screened out (|log_2_FC| > 1 and FDR < 0.05; FC: fold change), including 704 up-regulated miRNAs and 6 down-regulated miRNAs (Table S3). Among them, has-miR-1246 and has-miR-1307-3p, which were significantly overexpressed in breast cancer patients, have been proven to be potent combined markers for early detection of breast cancer in published studies, with a sensitivity of 97.3%, a specificity of 82.9%, and an accuracy of 89.7% ^18, 29^.

These 710 differential miRNAs were mapped to 11,418 target genes in the miRTarBase database (Table S4). As shown in Figure 1C, the results of Kyoto encyclopedia of genes and genomes (KEGG) pathway enrichment analysis revealed that proteoglycans in cancer, hippo signaling pathway and signaling pathway regulating pluripotency of stem cells, etc. were regulated by differential miRNAs and might participate in the onset and progression of cancer. In particular, these target genes were also significantly enriched in breast cancer pathway (FDR < 0.001; Table S5), which was consistent with the results of gene set enrichment analysis (GSEA) (enrichment score = 0.758, FDR < 0.001; Figure 1D, Table S6).

### BioDecoder enabled precise diagnosis of breast cancer

By leveraging the prior biological knowledge, a miRNA-Gene-Module-Pathway-Disease biological decoding path (miBDP) was extracted from databases to characterize the biological process of miRNAs in the body (Figure 2A). Based upon miBDP, we constructed the miRNA bio-interpretable neural network model (BioDecoder) for breast cancer diagnosis. The 710 differential miRNAs were fed into BioDecoder as input, and then 11,418 targeted genes, 116 modules and 70 pathways from miRTarBase and KEGG were used as hidden layers for information extraction (Table S4), followed by a disease layer that outputs the probability of breast cancer (Figure 2A and Figure S1). For comparison, similar neural network architecture was used in FCN. However, different from FCN, each neuron in BioDecoder represented a specific biological entity, and the links between adjacent layers were partially connected according to the real biological relationship, rather than fully connected (Figure S1). Moreover, in view of the class imbalance issue in the discovery cohort, the synthetic minority oversampling technique (SMOTE) was performed to balance the sample size of the two classes (i.e., control and cancer), thereby improving model stability (Figure 2B, C).

**Figure 2.**
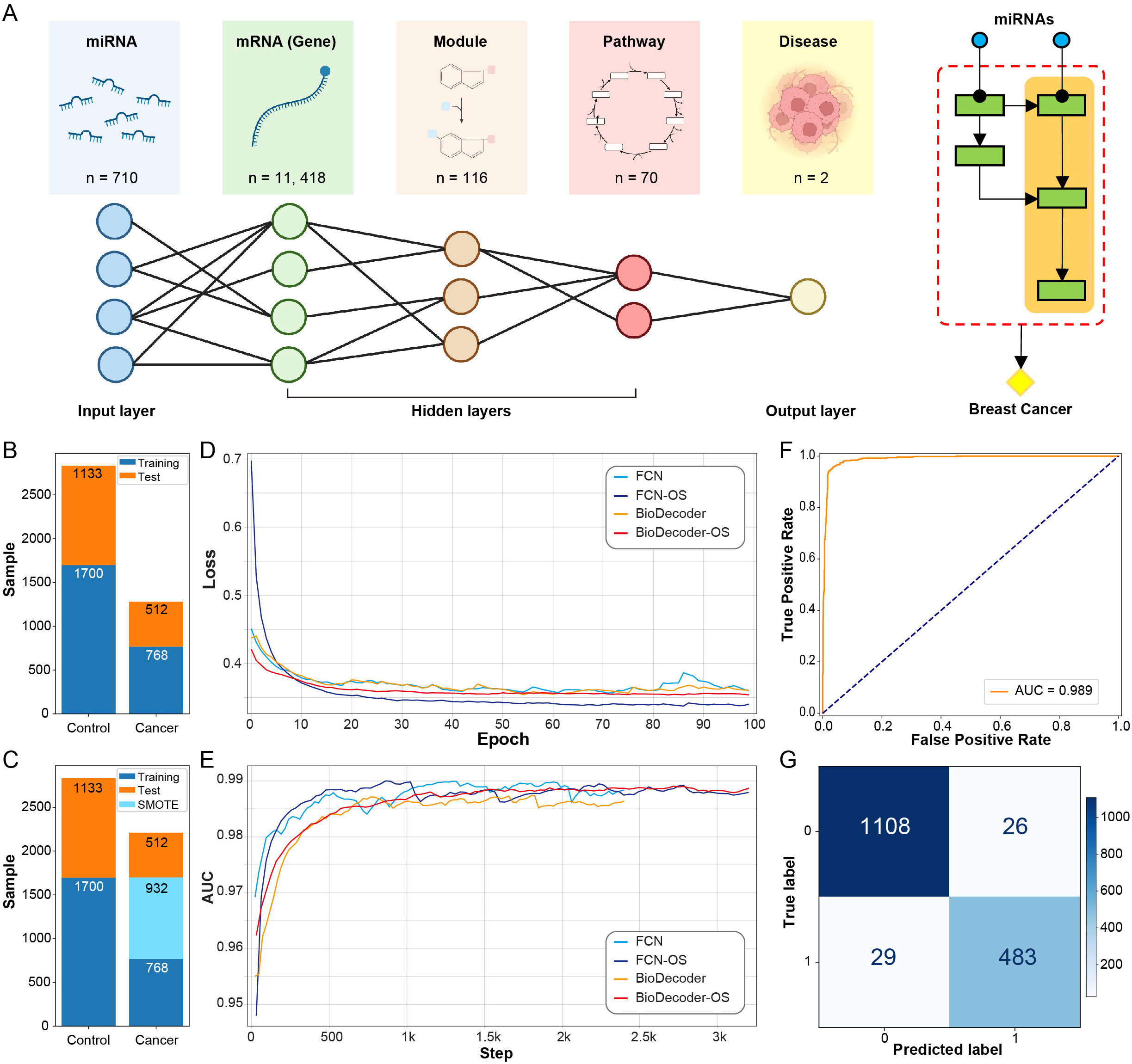
The architecture and performance of BioDecoder. (A) BioDecoder framework. This figure was created with BioRender.com (https://biorender.com/). (B) The distribution of control samples and breast cancer samples in the discovery cohort. (C) The distribution of control samples and breast cancer samples in the discovery cohort after oversampling. (D) The validation loss calculated by cross_entropy function during model training. (E) The AUC scores obtained during model training. (F) AUC of BioDecoder-OS on the test set. (G) Confusion matrix of BioDecoder-OS on the test set. SMOTE: synthetic minority oversampling technique; FCN: fully connected neural network; OS: oversampling; AUC: area under the receiver operating characteristic curve.

After 100 epochs training, the validation losses were minimized (Figure 2D) and the area under the receiver operating characteristic curves (AUC) was stable at the highest scores (Figure 2E). The results confirmed that RF, FCN and BioDecoder all had excellent prediction performance (AUC > 0.97; Table 1, Figure 2F and G, and Figure S2). Nevertheless, the validation AUC of RF was significantly higher than its test AUC, suggesting an overfitting problem. BioDecoder showed a comparable performance to FCN although it had more restrictions on model architecture (Table 1). BioDecoder with oversampling achieved the best performance for predicting risk of breast cancer on the test set (AUC = 0.989, balanced accuracy = 0.960, precision = 0.949, recall = 0.943) and was used for subsequent analysis.

**Table 1.** The predictive performance of random forest, fully connected neural network, and BioDecoder.

| Model | Training | Validation |  | Test |  |  |  |
| --- | --- | --- | --- | --- | --- | --- | --- |
|  | Loss | Loss | AUC | AUC | Accuracy* | Precision | Recall |
| **Raw data** |  |  |  |  |  |  |  |
| RF | NA | NA | 0.991 | 0.977 | 0.980 | 0.967 | 0.969 |
| FCN | 0.128 | 0.352 | 0.990 | 0.989 | 0.964 | 0.960 | 0.945 |
| BioDecoder | 0.889 | 0.359 | 0.988 | 0.986 | 0.955 | 0.948 | 0.934 |
| **Oversampled data** |  |  |  |  |  |  |  |
| RF | NA | NA | 0.995 | 0.977 | 0.979 | 0.963 | 0.971 |
| FCN | 0.125 | 0.348 | 0.989 | 0.988 | 0.958 | 0.951 | 0.938 |
| BioDecoder | 0.246 | 0.353 | 0.988 | 0.989 | 0.960 | 0.949 | 0.943 |

\* Balanced accuracy; RF: random forest; FCN: fully connected neural network; NA: not
applicable.

### BioDecoder revealed the underlying pathological mechanisms of miRNA in breast cancer

BioDecoder is a neural network architecture with bio-entity connections between adjacent layers (i.e., miRNA, gene, module and pathway), which can reflect the specific changes of these bio-entities in breast cancer. Ranking the pathways in BioDecoder by weights, it was found that several pathways, such as metabolic pathway, ribosome, oxidative phosphorylation, and DNA replication, were significantly different between the control and cancer samples (*P* < 0.001), and were of prime importance to breast cancer early diagnosis (Figure 3A, B).

**Figure 3.**
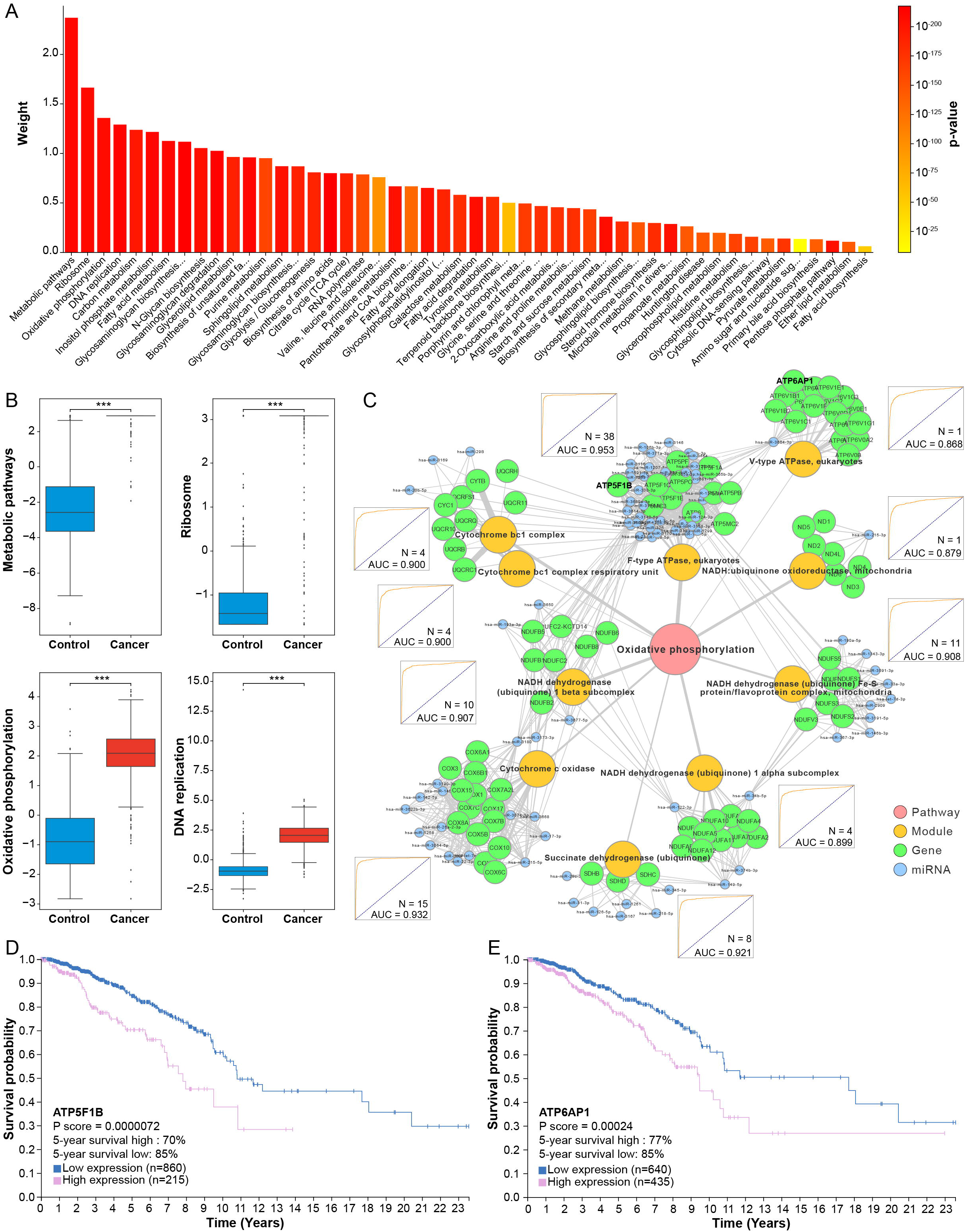
Biological interpretation of BioDecoder. (A) The pathway importance ranked by weights. (B) Boxplot of differential expression between control and breast cancer samples for the four important pathways (metabolic pathway, ribosome, oxidative phosphorylation, and DNA expression). (C) The biological network of oxidative phosphorylation pathway. Logistic regression was performed using miRNAs in each module and the receiver operating characteristic curves (ROC) were showed. (D). Survival curves of breast cancer patients based on ATP5F1B expression. Survival curves of breast cancer patients based on ATP6AP1 expression. ***,*P* < 0.001.

Specifically, hsa-miR-3659 and hsa-miR-190a-3p had high weights in the metabolic pathway (including 97 modules, 406 genes, and 403 miRNAs, Figure S3A) that has an important influence on breast cancer occurrence ^30^. The ribosome pathway is involved in the proliferation and metastasis of breast cancer cells ^31-35^, in which hsa-miR-17-3p and hsa-miR-3622b-3p were the key factors (Figure S3B). Oxidative phosphorylation contained 10 energy metabolism modules (e.g., F-type ATPase and V-type ATPase), 84 genes, and 81 miRNAs (Figure 3C). A subset of miRNAs targeting these modules also obtained good diagnostic capabilities for breast cancer. For instance, a set of 38 miRNAs (such as hsa-miR-3146 and hsa-miR-330-3p) in the F-type ATPase module achieved excellent diagnostic performance (AUC = 0.953), while the only miRNA (hsa-miR-3664-3p) in the V-type ATPase module yielded an AUC up to 0.868 (Figure 3C). Interestingly, we found that some target genes of miRNAs could affect the prognosis of breast cancer (Figure 3D, E). Low expression of ATP5F1B (*P* < 0.001) and ATP6AP137 (*P* < 0.001) significantly improved the breast cancer prognosis, and the 5-year survival increased from 70% and 77% to 85%, respectively.

### Extended application and validation of BioDecoder

The superior biological interpretability of BioDecoder opens up encouraging prospects in its clinical practice. As presented in Figure 4A, besides significantly distinguishing non-cancer and breast cancer samples (*P* = 2.666e-224), BioDecoder could also accurately identify other diseases, such as prostate disease (*P* = 2.302e-9) and benign breast disease (*P* = 1.539e-12). Meanwhile, although patients with benign breast disease were highly likely to develop cancer at miRNA level, the probability was still significantly lower than that of breast cancer patients (*P* = 0.040, Figure 4A). It indicated that BioDecoder has the potential for early screening of breast cancer.

**Figure 4.**
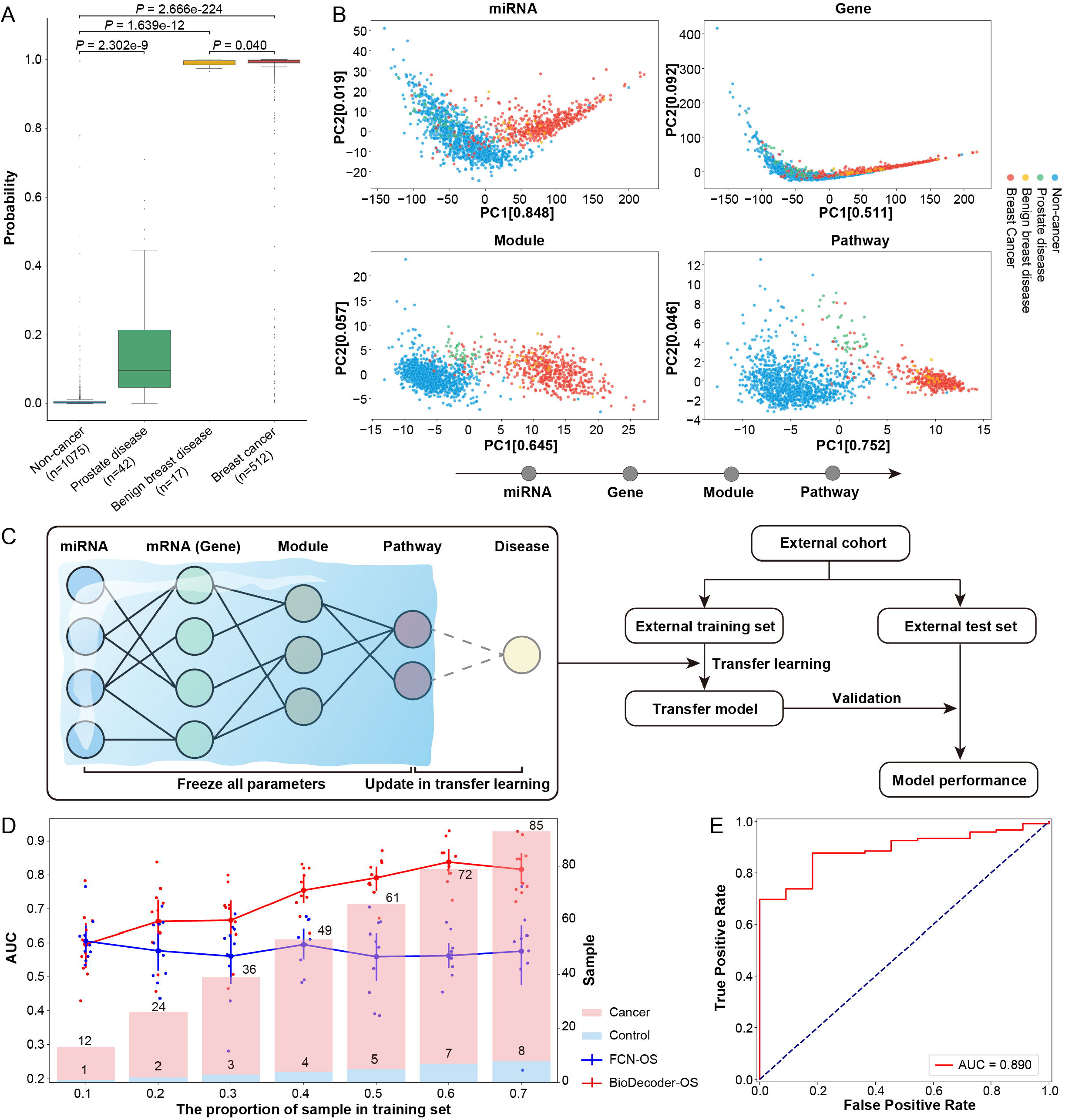
Application and validation of BioDecoder. (A) The probability of breast cancer predicted by BioDecoder in non-cancer, prostate disease, benign breast disease and breast cancer samples. The differences between groups were shown. (B) The distribution of test set samples at different miRNA biological decoding path levels of BioDecoder. (C) The flow chart of transfer learning for applying BioDecoder on the external cohort. (D) The transfer learning performance of BioDecoder and fully connected neural network on external cohort with different sampling proportions of the training set. (E) The receiver operating characteristic curve of BioDecoder’s transfer learning performance on the full external cohort. OS: oversampling.

BioDecoder contained the biological decoding path of miRNA, and the sample distribution of each level in miBDP is presented in Figure 4B. At the miRNA level, the model could roughly distinguish between control and breast cancer samples; nevertheless, disease samples were chaotic at principal component analysis (PCA) space. As the decoding proceeded, the distinctions between different categories increased gradually. At the module and pathway levels, there were significant differences among breast cancer, prostate disease and non-cancer samples, while benign breast disease samples were close to breast cancer samples (Figure 4B).

To evaluate the robustness and generalization ability of BioDecoder, an external validation was performed by transfer learning. The external validation cohort included miRNA expression profiles of breast tissue from 122 breast tumor patients and 11 healthy individuals ^36^. The first four layers of BioDecoder were frozen, and only the weights of the output layer were updated in transfer learning (Figure 4C). Taking into consideration different sampling proportions (10%–70%) of the external training set, BioDecoder exhibited better generalizability than FCN—BioDecoder achieved an excellent diagnostic performance with only a few external training samples, yielding AUC up to 0.890 (Figure 4D, E).

## Discussion

Breast cancer is the most common malignant cancer in women and its early diagnosis can effectively reduce mortality ^37^. The accuracy of breast cancer early screening has always been the coalface of research ^2, 38^. In this study, we designed a miRNA bio-interpretable neural network model, BioDecoder, for noninvasive diagnosis of breast cancer. Based upon miRNA expression profile of serum sample, BioDecoder achieved superior predictive performance for breast cancer (area under the receiver operating characteristic curve [AUC] = 0.989). In addition, BioDecoder showed strong robustness and clinical generalizability (AUC = 0.890) through transfer learning, even for breast tissue samples.

Liquid biopsy has been commonly used in cancer diagnosis due to its high sensitivity and specificity, especially with the aid of artificial intelligence (AI) ^39, 40^. Such black-box models, however, could hardly integrate into daily clinical practice owing to their poor bio-interpretability ^22^. To alleviate this issue, here we developed BioDecoder based on the architecture of miRNA biological decoding path (miBDP, miRNA-Gene-Module-Pathway-Disease) ^24, 41^. The bio-interpretable miBDP architecture not merely considerably reduces the number of parameters and enhances modeling efficiency (Figure S1) ^26^, but also guarantees BioDecoder great benefits in digging into the pathogenesis of breast cancer and discovering potential therapeutic targets (Figure 2, 3). The construction of Biodecoder is a decoding process of miRNA expression information according to miBDP, in which different diseases and their stages can be effectively distinguished (Figure 4B). Besides the excellent predictive power, BioDecoder exhibited outstanding performance in transfer learning (Figure 4D), implying that biologically interpretable architecture has an edge in terms of model generalizability and clinical application.

Clinically, circulating miRNAs has been proved to be related to the pathogenesis of breast cancer ^28, 42^, and can be used as biomarkers for breast cancer diagnosis, such as has-miR-1246 and has-miR-1307-3p ^18, 29^. In our results, 710 differential miRNAs were significantly enriched in breast cancer related pathways. Through the biological interpretation of miBDP, metabolism and oxidative phosphorylation were found to be the key pathways for miRNA to regulate the development of breast cancer, which indicates that metabolism may be reprogrammed in breast cancer ^27, 43, 44^. Moreover, under the regulation of miRNAs, some genes, such as ATP5F1B and ATP6AP1, could affect the prognosis of breast cancer. Studies have shown that the increased expression of genes in oxidative phosphorylation pathway plays an major role in the immunotherapeutic drug resistance of breast cancer, which could be reversed by the knockdown or inhibition of ATP synthase ^45^. Our findings suggests that BioDecoder’s interpretability can offer new thoughts for refining clinical diagnosis and precise treatment of breast cancer.

Although BioDecoder uncovered the key pathways for the onset and progression of breast cancer, the mechanism of miRNA targeting these pathways still needs experimental verification. In essence, the interpretability of BioDecoder comes from prior biological knowledge, and therefore detailed biological knowledge (e.g., genetic information, clinical characteristics, and various molecular experimental data) can improve model performance in capturing the real causality. Also, the transferability and predictive performance of similar architecture applied to other biomarkers and diseases need to be further evaluated.

## Conclusions

Our study proposed a bio-interpretable neural network architecture, namely BioDecoder, which can accurately diagnose breast cancer and reveal the potential mechanism of miRNA in breast cancer. Based on reliable prior knowledge, this bio-interpretable architecture has great potential to be applied to other types of biomarkers and diseases.

## Methods

### Discovery cohort

The data used in this work can be acquired from the ArrayExpress database (https://www.ebi.ac.uk/biostudies/arrayexpress). The discovery cohort (E-GEOD-73002, Table S1) consists of 1280 serum samples from breast cancer patients and 2833 serum samples from control samples (i.e., 2686 non-cancer samples, 93 prostate disease samples and 54 benign breast disease samples) ^18^. Samples from breast cancer patients with the following characteristics were excluded: (1) given drugs before serum collection and (2) with concurrent or previously diagnosed advanced cancer in other organs. Serum samples of control samples with no history of cancer or hospitalization within the past 3 months were included for analysis. The miRNA expression profiles of all samples were obtained by microarray analysis and verified by quantitative Reverse Transcription-Polymerase Chain Reaction (RT-PCR).

### External validation cohort

The external cohort includes 133 Spanish breast tissue samples (i.e., 122 breast cancer samples and 11 control samples, Table S2), which was reported by Matamala et. al. (https://www.ncbi.nlm.nih.gov/geo/query/acc.cgi?acc=GSE58606) ^36^. The miRNA expression profiles of all tissue samples were obtained by microarray analysis and verified by quantitative RT-PCR.

### Experimental setup

A stratified random sampling was performed to divide the discovery cohort into two subsets: 60% for training set (768 breast cancer samples and 1700 control samples) and 40% for test set (512 breast cancer samples and 1133 control samples). Then the training set was oversampled to balance the number of positive and negative samples using the synthetic minority oversampling technique (SMOTE) algorithm of the *imblearn* package (version 0.9.1) ^46^.

### Construction of artificial intelligence (AI) models

#### Random Forest (RF) model

The RF model was constructed by the *scikit-learn* (version 0.21.3) package. In the training set, we performed feature selection through recursive feature elimination using cross-validation. Subsequently, a 5-fold cross-validation and grid search were used for model training and hyperparameter tuning. Area under the receiver operating characteristic curve (AUC) was used as the primary evaluation measure for model selection. Finally, the RF model was constructed using the optimal features and hyperparameters (max_features = 0.1, n_estimators = 101, max_depth = None, max_samples = None, criterion = gini, and class_weight = balanced).

#### BioDecoder and Fully Connected Neural Network (FCN) models

The neural network models were constructed by the *pytorch* (version 1.13) package ^47^. The architecture of neural network consisted of one input layer (miRNA), three hidden layers (gene, module and pathway), and one output layer (disease). The input and hidden layers included linear function, rectified linear unit (ReLU) function, batch normalization (BatchNorm1d) function and dropout function, while the output layer contained only linear and BatchNorm1d functions, followed by the softmax function for classification (Figure S1). To make each layer biologically interpretable, we fixed the number of neurons according to the corresponding miRNA, gene, module and pathway, and links between adjacent layers were partially connected through a mask matrix, which was a boolean matrix representing real biological connections between layers, thereby providing biological meaning for the neurons between each layer. Notably, FCN had the same configuration as BioDecoder, except that the layers of FCN were fully connected and were not biologically meaningful (Figure S1).

The model was trained by Adam optimizer ^48^ (learning rate = 0.01, batch size = 64, and minimal epoch = 100) with batch gradient descent, and used cross entropy as the loss function. To prevent overfitting, the model was early stopped when the validation loss was minimized. The model was then applied to the test set to assess model performance. Evaluation metrics such as balanced accuracy, precision, recall and AUC were reported.

### Assessment of transfer learning robustness

Transfer learning ^49, 50^ was used to validate the predictive performance of BioDecoder on an external cohort. The first four layers of the BioDecoder were frozen, while the weights of pathway-disease were retrained by external training set (Figure S1). By the stratified random sampling, the external cohort was divided into two unequal parts— that is, the external training set and external test set. The training set was used to tune the transfer learning model (at the sampling proportion from 10% to 70%), and the test set was used to evaluate the model performance.

### Statistics Analysis

All statistical analysis was performed using *R* software (version 4.2.1) or *Python* software (version 3.9.6). Statistical significance was assessed using the Wilcoxon signed-rank test, unless otherwise specified. The differentially expressed miRNAs between breast cancer samples and control samples were established using a linear regression model in the R package *limma* ^51^. The resulting *P* values were corrected using the Benjamini-Hochberg (BH) method. The biomarkers that were differentially expressed miRNAs were screened by false discovery rate (FDR) < 0.05 and fold change (FC) > 2, or FDR < 0.05 and FC < 0.5. The corresponding target genes of differential miRNAs were obtained from miRTarBase database ^52^, and module and pathway information were extracted from the Kyoto encyclopedia of genes and genomes (KEGG). Pathway enrichment analysis was performed using the R packages *clusterProfiler* ^53^ and *GESA* ^54^. The PCA method from the python package *scikit-learn* was applied for principal component analysis (PCA). Gene expression data for breast cancer survival analysis were collected from the Human Protein Atlas (https://www.proteinatlas.org/). The network graph was visualized by *Cytoscape* (https://cytoscape.org/, version 3.9.0).

## Data Availability

No new sequencing data was used in this paper. All the software packages used in this study are open source and publicly available and the code used in this study is available on GitHub at https://github.com/ddhmed/BioDecoder.

https://github.com/tjcadd2020/BioDecoder

## Abbreviations

AI: Artificial Intelligence
AUC: Area Under the ROC Curve
BH: Benjamini-Hochberg
CTC: Circulating Tumor Cells
ctDNA: circulating tumor DNA
ctRNA: circulating tumor RNA
GSEA: Gene Set Enrichment Analysis
FC: Fold Change
FCN: Fully Connected Neural Network
FDR: False Discovery Rate
KEGG: Kyoto Encyclopedia of Genes and Genomes
miBDP: miRNA Biological Decoding Path
OS: Oversampling
miBDP: miRNA Biological Decoding Path
RF: random forest
ROC: Receiver Operating Characteristic
RT-PCR: Reverse Transcription-Polymerase Chain Reaction
SMOTE: Synthetic Minority Oversampling Technique
XAI: eXplainable Artificial Intelligence

## Key Points

- Artificial intelligence technology combines prior biological knowledge greatly improves the model interpretability while ensuring the prediction performance.
- BioDecoder achieved accurate early diagnosis of breast cancer and showed strong robustness and clinical expandability.
- The pathways, such as metabolic, ribosome, oxidative phosphorylation and DNA replication, played key roles in the pathogenesis of breast cancer.

## Acknowledgements

We are grateful for all the subjects who participated in this study. BioDecoder framework and its biological prior knowledge schematic were created with *BioRender* (https://biorender.com/), and network graph were draw with *cytoscape*.

## Authors’ contributions

Dingfeng Wu and Ruixin Zhu conceived and designed the project. Each author has contributed significantly to the submitted work. Lei Liu, Suqi Cao and Liu Yang was responsible for the data analysis and drafted the manuscript. Lixin Zhu, Sheng Gao, Weili Lin, Na Jiao, RuixinZhu and Dingfeng Wu revised the manuscript. All authors read and approved the final manuscript.

## Ethics approval and consent to participate

N/A

## Consent for publication

Obtained.

## Supplementary material

Figure S1. The neural network architecture of fully connected neural network, BioDecoder and transfer learning.

Figure S2. The receiver operating characteristic curve and confusion matrix of random forest, fully connected neural network and Biodecoder in raw data and oversampling data.

Figure S3. The biological network of metabolic pathway and ribosome pathway.

Table S1. Discovery cohort (E-GEOD-73002).

Table S2. The external validation cohort (GSE58606).

Table S3. The 710 miRNAs with significant differences between breast cancer and control samples.

Table S4. The correspondence of biological entries in miBDP.

Table S5. Pathway enrichment of miRNA targeted genes by clusterProfiler.

Table S6. Pathway enrichment of miRNA targeted genes by GSEA.

