## Supplementary figures and images for "BioDecoder: A miRNA Bio-interpretable Neural Network Model for Noninvasive Diagnosis of Breast Cancer"

### Figure S1

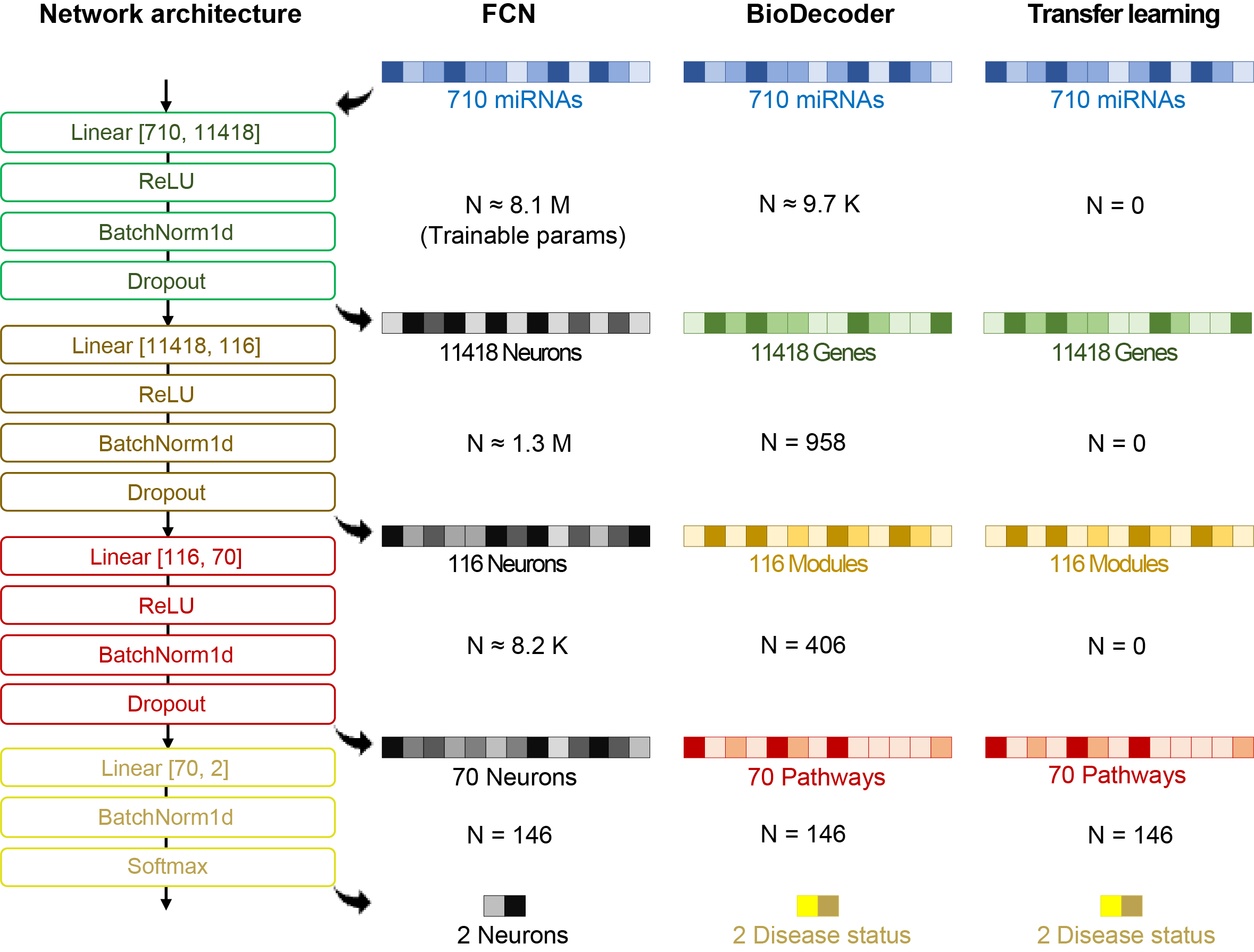

### Figure S2

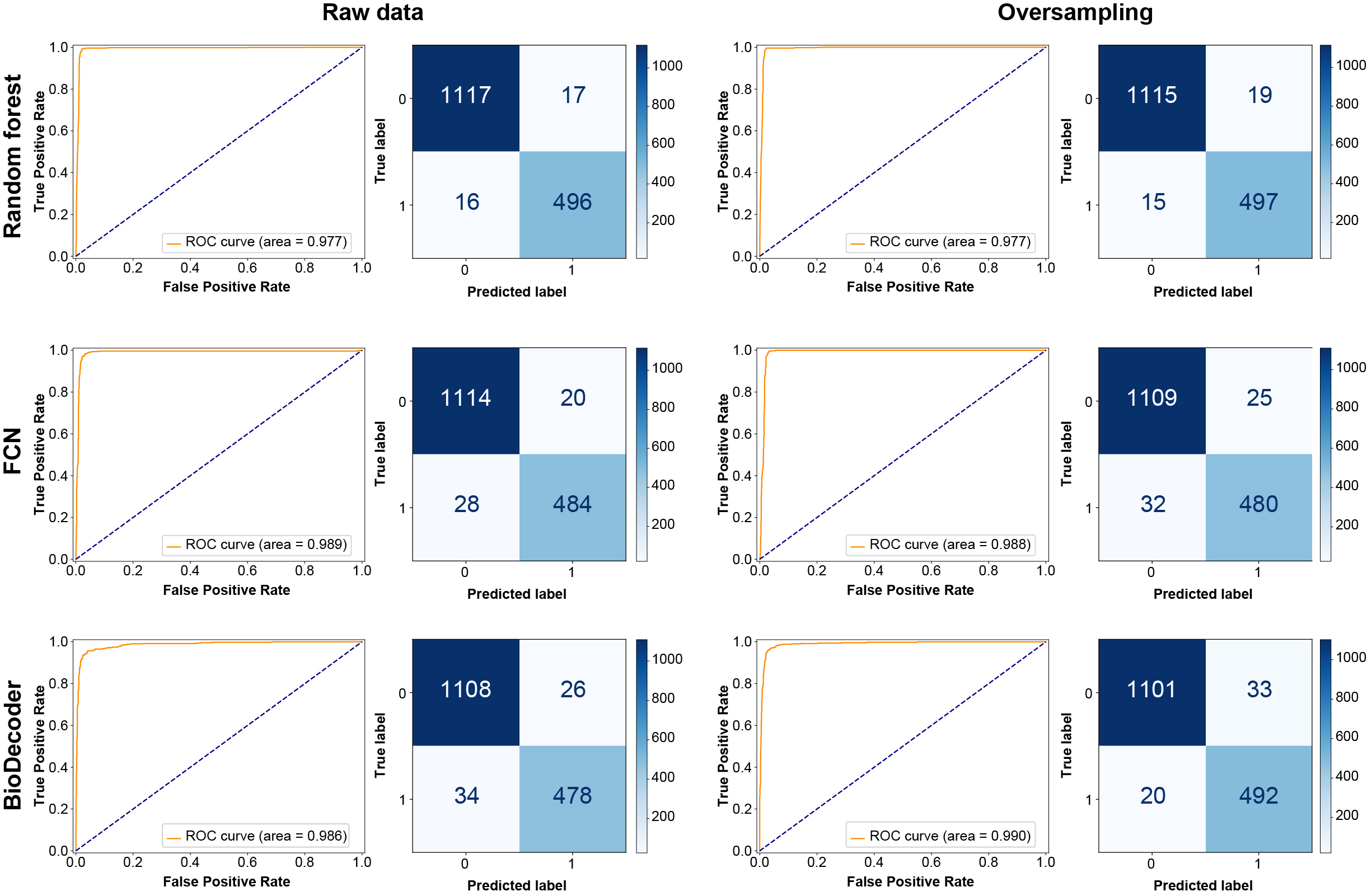

### Figure S3

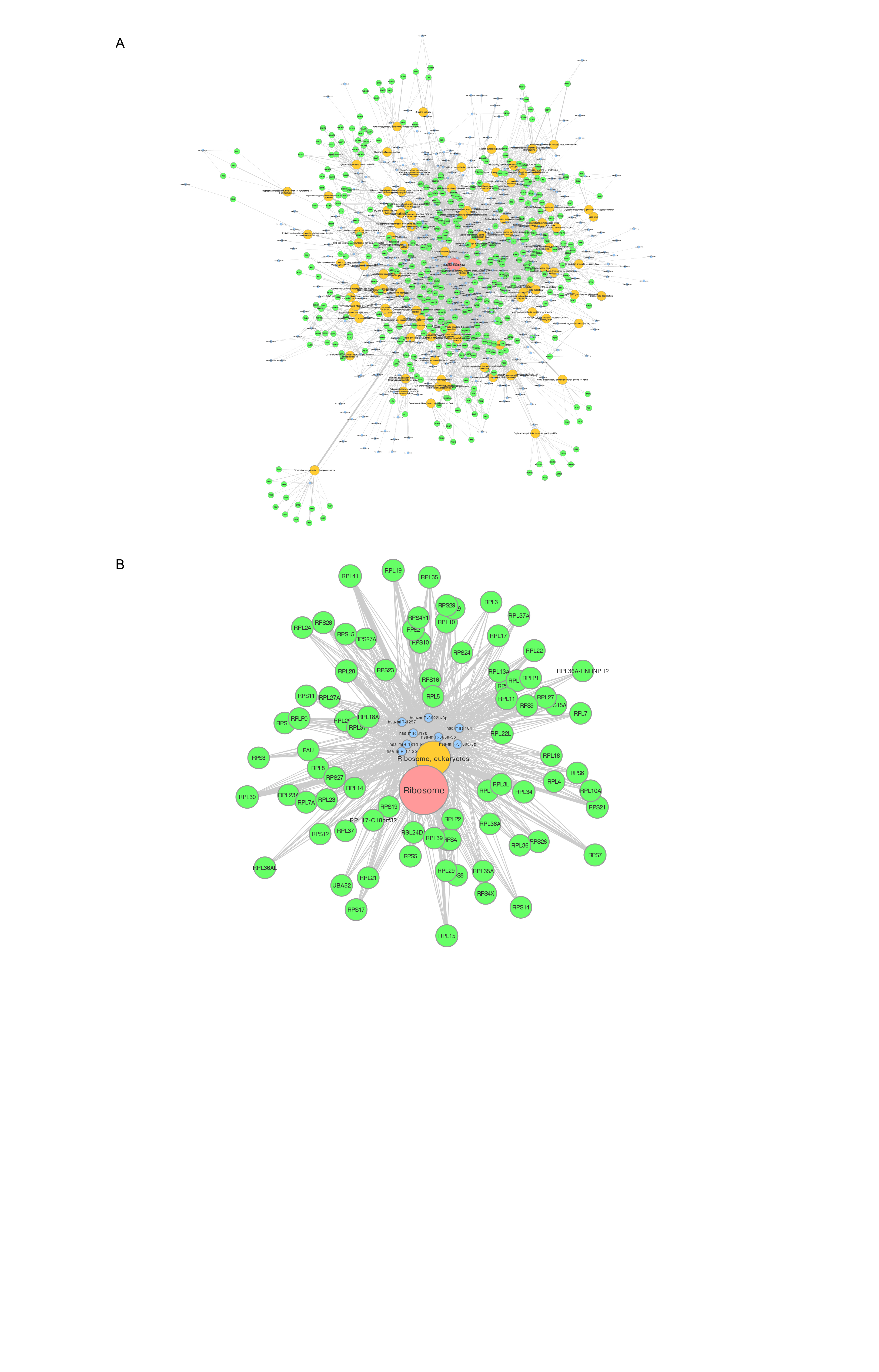
